# Knowledge of behavioural risk factors for types 2 diabetes mellitus and its associated factors among reproductive-age women in Arba Minch town, Gamo zone, 2022

**DOI:** 10.1101/2022.09.23.22280282

**Authors:** Tinsae Seyoum, Selamnesh Tesfaye, Yohannes Shiferaw, Rahel Hailu, Dagim Tefera, zeleke Gebru

## Abstract

**Background:** Type 2 diabetes accounts for over 90% of all types of diabetes. It is caused by a combination of behavioral risk factors. Currently, it is a serious health problem, especially in reproductive-age women associated with reproductive disorders. To prevent it, knowledge is vital, but there is a scarcity of data on behavioral risk factors in Ethiopia.

**Objective:** To assess knowledge of the behavioral risks of Type 2 diabetes mellitus and its associated factors among reproductive-age women in Gamo zone, Arba Minch town, 2022.

**Materials and Methods:** A community-based cross-sectional study was employed, and all women in the town were considered as the source population. A multistage sampling procedure with simple random sampling was adopted for the recruitment of kebeles. A systematic random selection procedure was also applied to the household with a 13th interval. A total of 623 samples were completed with an interview questionnaire. A bivariate logistic model was used to calculate the crude odds ratio, and multivariate analysis to control for confounding and identify the association for model fitting variables with AOR.

**Result:** The level of BRF knowledge among reproductive women is 47.0% [95% CI, 43.5-50.9], with the following factors having significant associations: average family income 3000–5000 Eth. Birr 1.81 [95% CI, 1.03-3.18], >= 5001 Eth. Birr 1.93 [95% CI, 1.02-3.68], DM in the friend or relatives 4.03[95% CI,1.56-10.46], Family history of DM 9.47 [95% CI, 4.74–18.90], source of information: health workers 1.87 [95% CI, 1.04-3.34] and friend or relatives 1.65 [95%CI,1.04-2.62].

**Conclusion:** The knowledge of behavioral risk factors for type 2 diabetes was poor among study participants. Family income, DM in the friend or relatives, family history of DM, and source of information were the associated factors with good knowledge. Health education about behavioral risk factors should be given emphasis broadly for women.

## Introduction

Diabetes mellitus (DM) is one of the main types of non-communicable disease caused by a combination of different factor (1, 2). Type 2 diabetes(T2DM) is a non-insulin-dependent diabetes resulted from an insufficient production of insulin and peripheral resistance to insulin (3). It accounts over 90% of all types of diabetes in the world and usually occurs in middle-aged and older people; however, it progressively affects in the most productive age group (4). People with diabetes have a high risk of developing serious health problems that leading to reduced quality of life (5). The high blood glucose levels disrupt different vital parts of the bodies including the heart, kidneys, eyes and nerves ending up various vascular complications (6). Besides to this, the progress of type 2 diabetes during the reproductive age group may rise the inter-generational risk (7). It is also identified as one of a challenge for sustainable development (1).

T2DM is evolving rapidly across the globe as a result of growing age, economic development, and urbanization, all of which result in behavioral risk factors. The incidence is rises gradually at puberty, likely due to hormonal changes and insulin resistance and it is also higher in female than men (4). On the other hand, it is linked to reproductive problems in women of reproductive age, which may be resolved with behavioral changes (7).

Around 451 million individuals over the age of 18 have diabetes and nearly 22 million women’s live birth are affected by the prenatal period (8). Diabetes affects fourteen percent of all live births in the Middle East and Africa (9). At the population level, diabetes prevention relies profoundly on public awareness of risk factors and appropriate risk perception (10).

Diabetes is caused by numerous reasons. Non-modifiable (family history of diabetes and age) as well as modifiable risk factors for type 2 diabetes exist. The behavioral risk factors (BRF) are modifiable factors that can increase the chance of developing diabetes mellitus; they include: obesity, physical inactivity, consuming an unhealthy diet, use of heavy alcohol, tobacco, sleeplessness, and stress (11, 12). These problems are hidden in low socio-economic status countries, including our country (13, 14).

BRF is one of the most important predisposing variables for T2DM. Conversely, the adoption of good behavioral changes and the reduction of negative lifestyles are an indication of knowledge (15). Knowledge of BRF is also a prerequisite for preventing diabetes in reproductive age groups. As a result, the goal of this study is to gain a better understanding of RA women’s knowledge of BRF for type 2DM and associated factors in Arba Minch, Southern Ethiopia. The findings of this study will also serve as a baseline for future intervention programs promoting early detection and control of diabetes risk factors.However, there is no available literature about the knowledge of BRF among WRA in Ethiopia. Therefore, this study will somehow fill a gap by assessing the level of knowledge about the behavioral risks of type 2 diabetes mellitus and its association among reproductive-age women who live in the Gamo zone, Arba Minch town.

## Methods and Materials

A community Based cross-sectional approach was employed from he study period from February 20/2022 to March 22, 2022, Arba Minch town.

The Source population were all Reproductive age women living in the Arba Minch town and the study population were all randomly selected Reproductive age women who were living in selected kebeles of Arba Minch town. Women reproductive ages that are living in Arba Minch town for at least six months were included in the study and Women who are severely ill, unable to respond during data collection time, and mentally ill were excluded.

To address the study, a multi-stage sampling strategy was utilized. Four kebele out of eleven in the town were randomly selected using a sample random sample technique, and the desired sample size was proportionally apportioned on the number of households to choose the kebeles in the survey.

Prior to data collection, a census was undertaken to identify the eligible population in the cho sen kebele.

The sample size determination was calculated using the formula of single population formula:

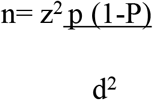

Where n = the desired sample size, Z=standard deviation at 95% confidence level (1.96),

P= is the proportion in the target population estimated to have a particular characteristic. P is, therefore, the proportion of knowledge of risk factors for DM in Nigeria was 75.5 % (16).

Since the study, q = proportion of people= 1-0.755 = 0.245 while it is the allowable error margin (d) of 5% = 0.05.

Therefore,

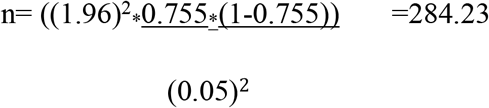

Using this formula, the sample size was calculated to be 284 and this multiplied by the design effect of 2 with the addition of 10% of non-response and the final sample size becomes **625**.

The computed sample size was proportional to the size of the population in each kebele after the number of households was obtained from the census.

The data was then collected by ten data collectors and four supervisors from house to house after three-day training. Then, using a systematic random sample approach, the study unit was chosen. For those randomly selected reproductive age women, a face-to-face interview was done to allow for the clarification of any questions. Each interview was last about 25 minutes. All questions were translated from English to Amharic and then back-translated to assure correctness before the study began, and a pretest was conducted with 5% of the sample population who lived outside the study area. The questionnaire’s validity and consistency were tested, and small changes were made based on the pretest.

Knowledge of BRF is the dependent variable of the study and the independent variables include; socio-demography variables, other diabetes related factors (relative and or friend history of DM, having diabetes in family, previously diagnosed, exposure to DM health education, heard about DM and source of information).

## Operational definitions

### Behavioural Risk Factors

Factors that increase the possibility of type 2 DM and can be modifiable and controllable: such as; physical inactivity, unhealthy diet, harmful use of alcohol, smoking, obesity, and stress (11, 16, 17).

### Knowledge of behavioural risks

Individuals who responded to behavioural risk-related questions: those answered below the mean value will be considered as Poor knowledge and Individuals who responded is ≥ the mean value as good knowledge (15, 17, 18).

### Data Collection

All questions were translated from English into Amharic and then back-translated to ensure accuracy. The study instrument was included questions on demographics, knowledge of behavioural risk factors (Physical activity, obesity, diet, tobacco use, stress, and sleep time). Standardized training to the attending data collectors was also given. Data collectors were degree holders of health professionals. The questionnaires were pre-validated with a pre-test of 5 % of the sample population out of the study area. The interviewed questionaries’ were offered to the selected WRA. The questionnaire’s internal consistency was assessed by measuring the Cronbach alpha coefficient for questionnaire scales and the test results were 0.79 (*r=0*.*787*).

A face-to-face interview was conducted to open opportunities for clarifying any questions at a suitable time. The estimated time for each interview was around 25 minutes.

### Data quality management

For data quality measures: training of data collectors on their duties for the purposes of the study and supervisors, pre-test on 5 % sample on non-selected area to avoid information contamination bias, some modification was done and a daily report to the supervisors and investigators were strictly performed to ensure the completeness of the data and to offer additional clarification. To avoid data entry error, double data entry through Epi-data was used with proper labeling and coding of data was done during data cleaning stages.

### Data processing and Analysis

To enter data, Epi Data 4.6 was used, and SPSS version 21 was used for all statistical analyses. Data editing was carried out by checking and verifying the completed questionnaire at the end of the interview, as well as at the end of the whole survey and before the analysis. A descriptive analysis was conducted to describe the distribution of the demographic factors and the level of knowledge of the behavioral risk of T2DM. Each correct response was given a score of 1, and a score of 0 for wrong or uncertain responses. Each level of the BRF is given a separate score for twelve questions, which is then added together for a maximum score of twelve (12). A mean score of less than 4.60 was poor knowledge and 4.60 and above was good knowledge of BRF for T2DM.

Binary logistic regression analysis was conducted to assess the association between the independent with the dependent variables and to identify determinants of behavioural risk factors for T2DM. Crude odds ratio (COR) and adjusted odds ratio (AOR) with its respective 95% confidence interval (CI) was used to interpret the result. Then, multivariable logistic regression was used to control the possible confounders on a binary logistic regression, with a confidence interval of 95%, P-value < 0.05. A Descriptive statistic was used to describe the study participants in relation to relevant variables. Then, bivariate and multivariate logistic regression was used to identify the possible associations between independent and outcome variables. Variables with p-values less than 0.25 during bivariate analysis were transferred to multivariable analysis to control for confounding effects. Variables with a p-value of < 0.05 in the multivariable analysis were considered significantly associated with the outcome variable. Assumption tests and model goodness of fit were checked. The goodness of fitness of the model was checked by Hosmer and Lemeshow test (p value =0.160). A Multi-Collinearity test between predictable variables was checked by using the Variance inflation factor with cut off value < 5 and tolerance > 0.1, and correlations were tolerable. Finally, the results of the study are presented in tables, figures, and text.

### Ethical Considerations

This study was approved by the Arba Minch College of health science review board. An ethical clearance letter (Ref No/አ/ም/ጤ/ሳ/ኮ/01/ 20/353) was obtained from the Ethical Review Committee of the AMCHS. A Letter from the Research Ethics Committee was submitted to respective offices. During data collection, informed verbal and written consent was obtained from respondents and ascent from family after explaining the purpose of the study. To minimize the transmission of COVID 19 as much as possible, the data collectors were strictly followed the procedure.

## Results

### Socio-demographic characteristics of the respondents

Of the total of 625 calculated study participants, 623 were agreed to participate in the study that yielding a response rate of 99.68%. The minimum age of respondents is 15-year-old and maximum age is 49 years. The mean age of respondents was 27.3 (SD+8.2 years). Majority respondents were married (57.6%). Concerning the educational status, 27 (4.3%) was illiterate, and able to write and read and majority 235 (37.7%) were collage and above. One hundred eighty-one (29%) were students in occupation. The average family income was 3910.00 Eth birr (See table1).

**Table 1.** Socio-demographic status of respondents on Behavioural Risk Factors of Type -2-Diabetes Mellitus in Arba Minch town 2022 [N=623].

| Variables |  | Frequency | Percentage | CI, 95% |
| --- | --- | --- | --- | --- |
| Age | 15-24 year | 243 | 39 | 35.0-42.9 |
|  | 25-30 year | 177 | 28.4 | 24.6-31.9 |
|  | 31-40 year | 126 | 25.4 | 22.2-28.9 |
|  | >=41 year | 77 | 7.2 | 5.1-9.3 |
| Marital status | Single | 232 | 37.2 | (33.5 - 41.1) |
|  | Married | 359 | 57.6 | (53.7 - 61.4) |
|  | Separate/divorce | 16 | 2.6 | (1.6 - 4.1) |
|  | Divorce | 16 | 2.6 | (1.6 - 4.1) |
| Educational level | Illiterate | 27 | 4.3 | (3.0 - 6.2) |
|  | Read and write | 27 | 4.3 | (3.0 - 6.2) |
|  | Grade 1-4 | 35 | 5.6 | (4.2 - 7.9) |
|  | Grade 5-8 | 121 | 19.4 | (16.5 - 22.7) |
|  | Grade 9-12 | 178 | 28.6 | (25.2 - 32.3) |
|  | Collage and above | 235 | 37.7 | (34.0 - 41.6) |
| Occupation | House wife | 164 | 26.3 | (23.0 - 29.9) |
|  | Student | 183 | 29.1 | (25.7 - 32.8) |
|  | Governmental employee | 128 | 20.5 | (17.6 - 25.9) |
|  | Merchant | 84 | 13.5 | (11.0 - 16.4) |
|  | Private employee | 26 | 4.2 | (2.9 - 6.0) |
|  | Daily labor | 19 | 3 | (2.0 - 4.7) |
|  | unemployed | 19 | 3 | (2.0 - 4.7) |

#### 5.1 Awareness of participants on DM

Regarding awareness, almost 90.5% of study participants had heard about DM and 264 (42.2%) had gained information from friends or relatives. From the total study participants, 159 (25.4%) had received health education about DM and those who had friends or relatives with diabetes were 192 (31%); besides this, 134 (21.5%) respondents had had diabetes in their families (Table 2).

**Table 2.** The description of response of study participant on the socio-economic and awareness status in Arba Minch, 2022 (n=623).

| **Variables** | **Response** | **No** | **percent** | **95%CI** |
| --- | --- | --- | --- | --- |
| Monthly Average family income | <1999 Eth. birr | 161 | 25.8 | 22.6-29.1 |
|  | 2000-2999Eth. birr | 91 | 14.6 | 34.5-41.9 |
|  | 3000-5000 Eth. Birr | 232 | 13.5 | 10.9-16.4 |
|  | >50001 Eth. Birr | 139 | 22.3 | 19.1-25.7 |
| Heard about DM | Yes | 568 | 91.2 | (88.0 - 92.6) |
|  | No | 55 | 8.8 | (7.4 - 12.0) |
| Source of information<br>N=568 | Media | 185 | 32.5 | (26.1 - 33.2) |
|  | Health workers | 116 | 20.5 | (15.8 - 21.9) |
|  | Friend /relatives | 267 | 47 | (38.6 - 46.3) |
| Have got DM health education | Yes | 159 | 25.5 | (22.0 - 28.8) |
|  | No | 464 | 74.5 | (71.2 - 78.0) |
| Diabetes in friend /relatives | Yes | 192 | 30.8 | (27.3 - 34.6) |
|  | No | 395 | 63.4 | (59.5 - 67.1) |
|  | I don't know | 36 | 5.8 | (4.2 - 7.9) |
| Family with Diabetes | Yes | 134 | 21.5 | (18.5 - 24.9) |
|  | No | 458 | 73.5 | (69.9 - 76.8) |
|  | I don't know | 31 | 5 | (3.5 - 7.0) |

#### 5.2 Knowledge of Type-2DM

In respect of diabetes knowledge, in table 3, the majority of respondents (511, or 82.0%) didn’t know about T2DM that could result from insufficient use of insulin, and similarly, around 86.0% of participants didn’t distinguish that it could result from improper response of insulin. Of all the study participants, 39.8% knew that DM had a high level of sugar in the blood. Half of the participants (312, 50.1%) knew T2DM as incurable and 53.6% didn’t know about DM that could affect all parts of the body. The average percentage of correct responses regarding defining DM was 29%.

**Table 3.** The description of general knowledge about Type-2-DM definition among Reproductive age Women in Arba Minch, 2022.

| Variable | Yes |  | No |  | I don't know |  |
| --- | --- | --- | --- | --- | --- | --- |
|  | No | % | No | % | No | % |
| Define T2DM |  |  |  |  |  |  |
| T2DM is a condition of insufficient use of insulin | 57 | 9.10 | 55 | 8.80 | 511 | 82.00 |
| T2DM is a condition of the body which not responding for insulin | 58 | 9.30 | 30 | 4.80 | 535 | 85.90 |
| T2DM is a condition of high level of sugar in the blood | 248 | 39.80 | 41 | 6.60 | 334 | 53.60 |
| DM is not curable | 312 | 50.10 | 162 | 26.00 | 149 | 23.90 |
| DM is a disease which affect any part of the body | 230 | 36.90 | 59 | 9.50 | 334 | 53.60 |

##### 5.2.1 Knowledge of risk factors for T2DM

Among study participants, old age 345 (55.4%) and heredity (52.8%) were both mentioned as risk factors for T2DM by survey participants. However, these two factors are non-modifiable.

Regarding to the knowledge of BRF of diabetes, about half of respondents (354; 56.8%) and (351; 56.3%) identified excessive sugar and sweet foods, and poor dietary habits respectively. Around half of the participants were also stated as obesity 409 (49.3%) could be a risk factor and 275 (44.1%) reported that all T2DM risk factors could be preventable. Being hypertensive and not getting enough activity or exercise were also identified as risk factors for T2DM (42.4 percent and 41.3 percent, respectively). In addition to this, the lowest response indicated sedentary lifestyle (32.6 percent), insomnia (21.7%) and DM in pregnancy (gestational diabetes) (19.4 percent) as a risk factor (see table 4).

**Table 4.**
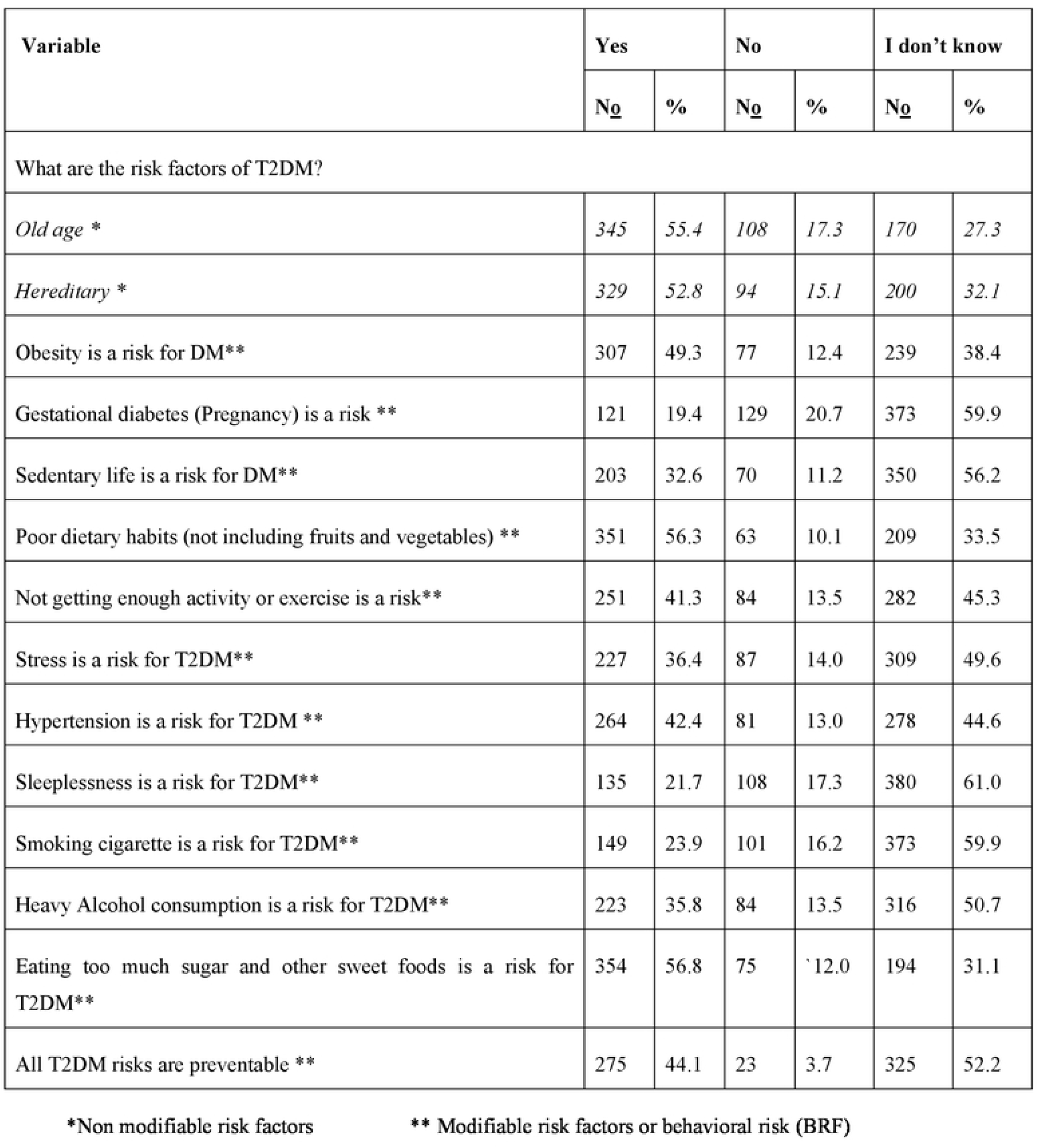
The Frequency distribution of participants response on knowledge of behavioural risk factors for type 2 diabetes, Arba Minch town, 2022 (n= 623).

| Variable | Yes |  | No |  | I don't know |  |
| --- | --- | --- | --- | --- | --- | --- |
|  | No | % | No | % | No | % |
| What are the risk factors of T2DM? |  |  |  |  |  |  |
| *Old age \** | 345 | 55.4 | 108 | 17.3 | 170 | 27.3 |
| *Hereditary \** | 329 | 52.8 | 94 | 15.1 | 200 | 32.1 |
| Obesity is a risk for DM** | 307 | 49.3 | 77 | 12.4 | 239 | 38.4 |
| Gestational diabetes (Pregnancy) is a risk ** | 121 | 19.4 | 129 | 20.7 | 373 | 59.9 |
| Sedentary life is a risk for DM** | 203 | 32.6 | 70 | 11.2 | 350 | 56.2 |
| Poor dietary habits (not including fruits and vegetables) ** | 351 | 56.3 | 63 | 10.1 | 209 | 33.5 |
| Not getting enough activity or exercise is a risk** | 251 | 41.3 | 84 | 13.5 | 282 | 45.3 |
| Stress is a risk for T2DM** | 227 | 36.4 | 87 | 14.0 | 309 | 49.6 |
| Hypertension is a risk for T2DM ** | 264 | 42.4 | 81 | 13.0 | 278 | 44.6 |
| Sleeplessness is a risk for T2DM** | 135 | 21.7 | 108 | 17.3 | 380 | 61.0 |
| Smoking cigarette is a risk for T2DM** | 149 | 23.9 | 101 | 16.2 | 373 | 59.9 |
| Heavy Alcohol consumption is a risk for T2DM** | 223 | 35.8 | 84 | 13.5 | 316 | 50.7 |
| Eating too much sugar and other sweet foods is a risk for T2DM** | 354 | 56.8 | 75 | 12.0 | 194 | 31.1 |
| All T2DM risks are preventable ** | 275 | 44.1 | 23 | 3.7 | 325 | 52.2 |

\*Non modifiable risk factors
\*\* Modifiable risk factors or behavioral risk (BRF)

The level of Knowledge of BRF for T2DM among reproductive age women was computed and the level of good knowledge was 47% (mean score ≥ 4. 60 with SD 3.46) (see fig.1).

**Fig1.**
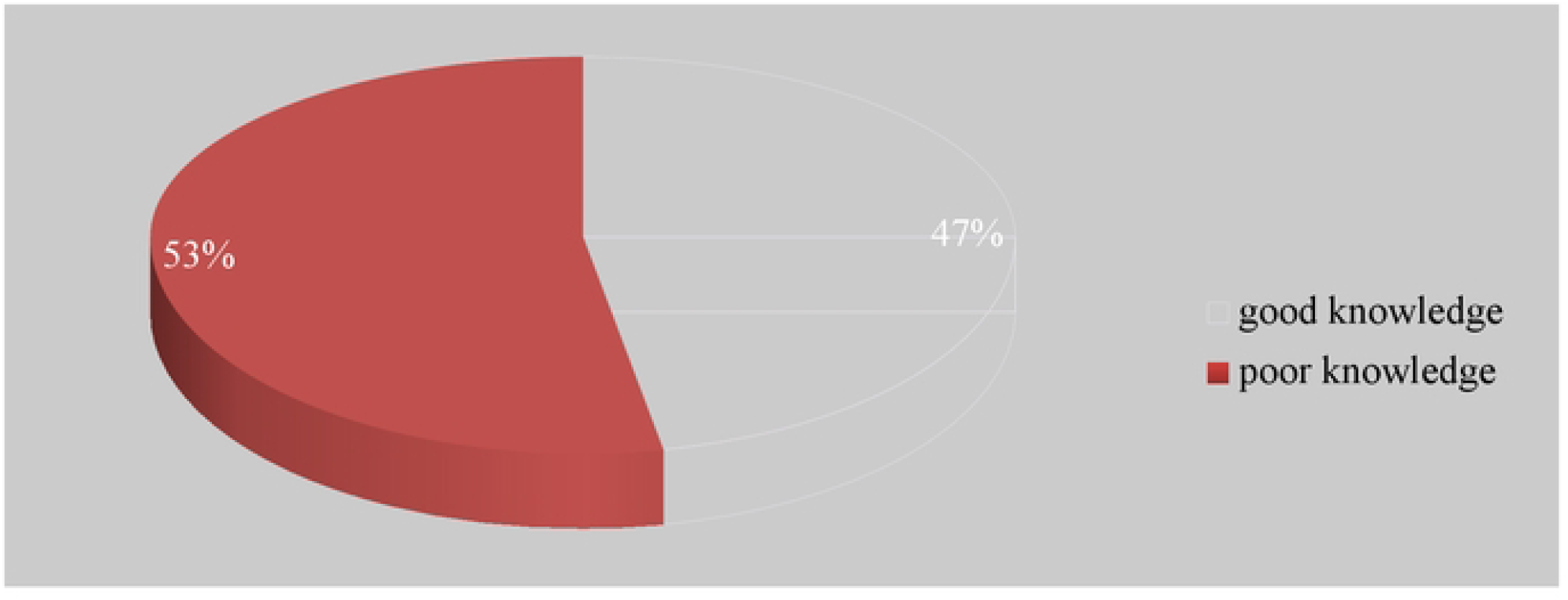
Knowledge of Behavioural Risk Factors for Type-2DM among reproductive age, women in Arba Minch town, 2022.

##### 5.2.2 Knowledge of symptoms of Type 2-DM

The most frequently mentioned symptom was excessive hunger 416 (66.8%) (See fig.2), followed by a high level of blood sugar (52%) and excessive thirst (50%), which was also reported as signs of diabetes. The least well-known symptom of DM among women of reproductive age was blurred vision, which accounted for 199 (31.9 %) of the total knowledge of symptoms among respondents.

**Fig 2.**
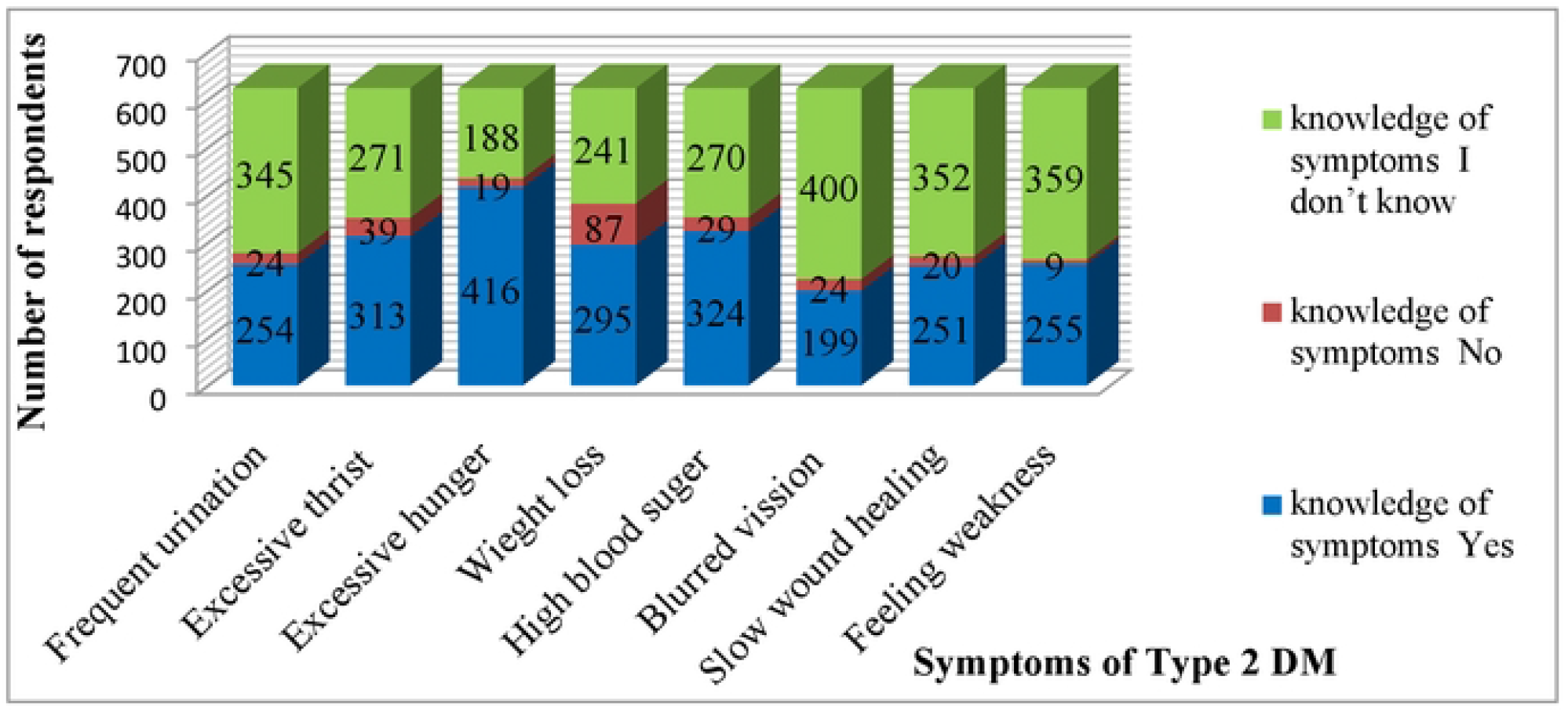
The response of reproductive age women on their knowledge of symptoms for T2DM in Arba Minch, 2022(N=623).

##### 5.2.3 Knowledge relate to prevention of DM and its complication

The responses to questions related to the prevention of diabetes complications are shown in table 5. Of all respondents, 458 (73.5%) were able to mention the prevention of diabetes complication using available medicines. Around half of the participants (51%) also knew that eating a healthy diet (more fruits and vegetables in serve) every day could help prevent DM, and 47 per cent said that proper weight reduction and exercising regularly could help prevent DM and its complications. The overall average of respondents’ knowledge of DM to prevent T2DM and its complications was 55%.

**Table 5.**
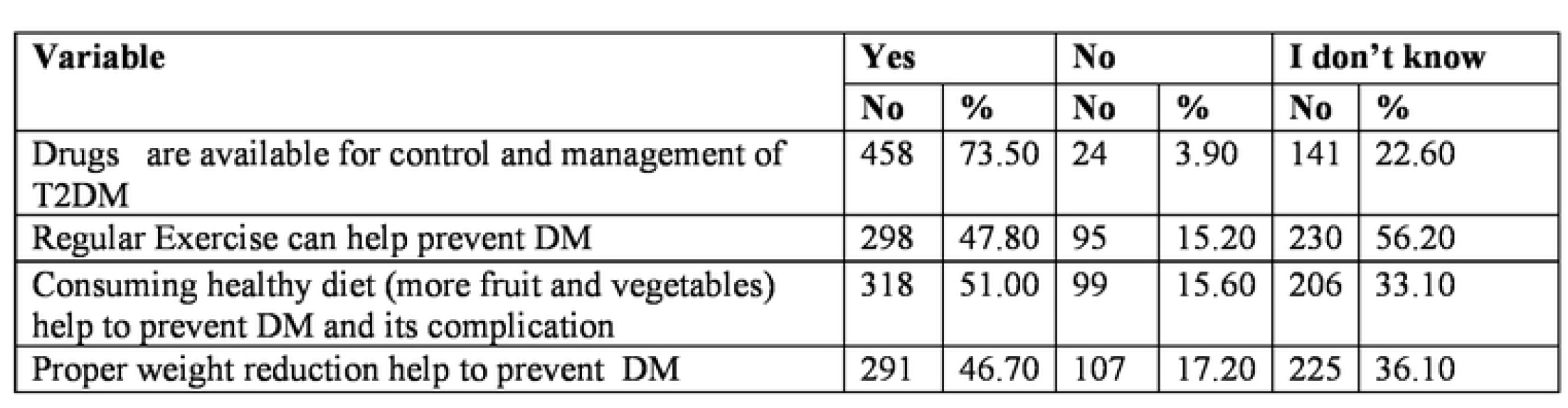
The knowledge of complication prevention of Type 2DM among Reproductive Age Women in Arba Minch town, 2022(n=623)

| Variable | Yes |  | No |  | I don't know |  |
| --- | --- | --- | --- | --- | --- | --- |
|  | No | % | No | % | No | % |
| Drugs are available for control and management of T2DM | 458 | 73.50 | 24 | 3.90 | 141 | 22.60 |
| Regular Exercise can help prevent DM | 298 | 47.80 | 95 | 15.20 | 230 | 56.20 |
| Consuming healthy diet (more fruit and vegetables) help to prevent DM and its complication | 318 | 51.00 | 99 | 15.60 | 206 | 33.10 |
| Proper weight reduction help to prevent DM | 291 | 46.70 | 107 | 17.20 | 225 | 36.10 |

#### 5.3 Behavioral risk Perception for Type 2DM

From 623 study participants, 68.7% of individuals agreed to be tested for diabetes, and 387 (61.1%) thought their family members should be checked as well. Avoiding fatty and sugary foods is thought to help control diabetes by 62.1 percent, while physical activity is thought to help prevent diabetes by 53.3 percent. Overall, 59 percent of respondents agreed with their risk assessment (see table 6).

**Table 6.** The perception of reproductive age women on behavioural risks of Type 2DM in Arba Minch town, 2022(n=623)

| Variable | Agreed |  | I Don't know |  | Disagreed |  |
| --- | --- | --- | --- | --- | --- | --- |
|  | No | % | No | % | No | % |
| Do you think that you should be examined for DM? | 428 | 68.70 | 142 | 22.80 | 53 | 8.50 |
| Do you think family members should be screened for DM? | 387 | 62.10 | 196 | 31.50 | 40 | 6.40 |
| Do you think should you follow avoiding of consumption of too much sugar to control DM? | 332 | 57.80 | 256 | 36.80 | 35 | 5.50 |
| Do you think physical activity can prevent the risk of DM? | 274 | 53.30 | 328 | 41.10 | 21 | 5.60 |
| Do you think maintaining a healthy weight is important in the management of diabetes? | 268 | 43 | 330 | 53 | 25 | 4 |
| Do you think early medical follow up can help to recognize DM and save life earlier? | 445 | 71.4 | 158 | 25.4 | 20 | 3.2 |

#### 5.4 Adopting healthy life style for Type 2 DM

Regarding adopting a healthy lifestyle, 235 (37.7%) of research participants adopted healthy lifestyles that promote diabetes impediment, which is critical for diabetes prevention. Only 7.9% of respondents regularly consume more fatty (saturated fat) and sweet (sugar) foods, which can contribute to diabetes, and 7.9% regularly exercise to maintain a healthy weight, but only 17 (2.7%) and 4.3 percent of respondents regularly monitor their blood sugar and blood pressure, respectively. The majority of individuals (477, or 76.6%) did not consume any alcohol, and almost all (621, or 99.7%) did not smoke cigarettes (See table 7).

**Table 7.** The description of behavioural risk and adoption of health status of Women Reproductive Age for Type 2DM in Arba Minch town 2022(n=623)

| **Variable** | **Frequently** |  | **Occasionally** |  | **Never** |  |
| --- | --- | --- | --- | --- | --- | --- |
|  | **No** | **%** | **No** | **%** | **No** | **%** |
| **Do you consume of fatty and sweets food?** | 49 | 7.9 | 472 | 75.8 | 102 | 16.4 |
| **Do you do 30-60 mints physical activity daily?**<br>**E.g. Brisk walking, house activities, climbing staircase.** | 188 | 30.2 | 314 | 50.4 | 121 | 19.4 |
| **Do you participate in maintaining your healthy weight?** | 49 | 7.9 | 379 | 61 | 195 | 31.3 |
| **Do you check your blood sugar regularly?** | 17 | 2.7 | 177 | 28.4 | 429 | 68.9 |
| **Do you check your blood pressure regularly?** | 27 | 4.3 | 225 | 36.1 | 371 | 59.6 |
| **Do you drink alcohol?** | 7 | 1.1 | 139 | 22.3 | 477 | 76.6 |
| **Do you smoke tobacco?** | 0 | 0 | 2 | 0.3 | 621 | 99.7 |

#### 5.5 Factors associated with knowledge of Behavioral Risk Factors for Type 2DM

In bivariate logistic regression analysis, age, educational status, occupation, average family income, source of information, family history of DM, and DM in their friends or relatives were significantly associated with BRF knowledge. Hence, these significant variables with a P value of less than 0.25 were entered into a multivariate logistic regression model for each potential predictor of knowledge of BRF of T2DM. Finally, age, educational status, and occupation were found to be non-significant. Analysis revealed that those with the average family income between 3000 and 5000 Eth. Birr was 1.81 times (AOR = 1.81, 95% CI = 1.03, 3.18) more likely had good knowledge about BRF than those with a monthly income of less than 1,999 Eth. Birr. A household with an average income of more than 5,001 Eth. Birr was also 1.93 times (AOR = 1.93, 95% CI = 1.02, 3.68) more likely to had good knowledge about BRF than its counterparts. Those having a friend or relative’s history of DM were 4.03 times (AOR = 4.03, 95% CI = 1.56, 10.47) more likely to had good knowledge about BRF than those who did not know having DM in their friends or relatives. Those having a family history of DM were 9.47 (AOR = 9.47, 95% CI = 4.74, 18.90) times more likely to know of BRF than those without a DM family history. Having information from health workers were 1.87 times (AOR = 1.87, 95 percent CI = 1.04, 3.34) have had good knowledge about BRF than with media, but having information from friends or relatives were 1.65 times (AOR = 1.65, 95 percent CI = 1.04, 2.62) likely to have had knowledge about BRF than those who got information from the media (See table 8).

**Table 8.** Bi-variable and multi-variable logistic regression predicting knowledge related to Behavioural Risk Factors for Type 2 Diabetes Mellitus among women reproductive age group of Gamo zone, Arba Minch towns, 2022 (N = 623).

| Variables | Categories | BRF knowledge level |  |  |  | COR [CI 95%] | P value | AOR [CI 95%] | P value |
| --- | --- | --- | --- | --- | --- | --- | --- | --- | --- |
|  |  | Good |  | Poor |  |  |  |  |  |
|  |  | No | % | No | % |  |  |  |  |
| Age | 15-24 | 99 | 15.9 | 144 | 23.1 | 1 |  | 1 |  |
|  | 25-30 | 78 | 12.5 | 99 | 15.9 | 1.15[0.77-1.69] | 0.495 | 0.58[0.31-1.07] | 0.079 |
|  | 31-40 | 88 | 14.1 | 70 | 11.2 | 1.83[1.22-2.74]* | 0.003 | 0.79[0.41-1.54] | 0.495 |
|  | >= 41 | 28 | 4.5 | 17 | 2.7 | 2.39[1.25-4.61]* | 0.009 | 0.61[0.24-1.57] | 0.306 |
| Average Family income | < 1999 | 52 | 8.3 | 109 | 17.5 | 1 |  | 1 |  |
|  | 2000-3000 | 74 | 6.9 | 85 | 7.7 | 1.83[1.16-2.87] | 0.009* | 1.59[0.81-3.14] | 0.174 |
|  | 3001-5000 | 84 | 18.5 | 80 | 18.8 | 2.20[1.40-3.45] | 0.001* | 1.81[1.03-3.18]** | 0.039 |
|  | >=5001 | 83 | 13.3 | 56 | 9 | 3.11[1.94-4.99]* | 0.000* | 1.93[1.02-3.68]** | 0.044 |
| Educational status | Illiterate | 11 | 1.8 | 16 | 2.6 | 1 |  | 1 |  |
|  | Read and write | 15 | 2.4 | 12 | 1.9 | 1.82[0.62-5.35] | 0.278 | 1.88[0.50-7.12] | 0.355 |
|  | Grade 1-4 | 15 | 2.4 | 20 | 3.2 | 1.09[0.39-3.02] | 0.867 | 1.02[0.28-3.72] | 0.975 |
|  | Grade 5-8 | 42 | 6.7 | 79 | 12.7 | 0.77[0.33-1.82] | 0.333 | 0.87[0.29-2.56] | 0.794 |
|  | Grade 9-12 | 72 | 11.6 | 106 | 17 | 0.99[0.43-2.25] | 0.977 | 1.05[0.36-3.06] | 0.933 |
|  | Collage & above | 138 | 23.2 | 97 | 15.6 | 2.07[0.92-4.65] | 0.079 | 1.43[0.46-4.39] | 0.537 |
| Occupation | House wife | 72 | 11.6 | 92 | 14.8 | 1 |  | 1 |  |
|  | Student | 76 | 12.2 | 107 | 17.2 | 0.91[0.59-1.39] | 0.656 | 0.74[0.37-1.50] | 0.408 |
|  | Merchant | 38 | 6.1 | 46 | 7.4 | 1.06[0.62-1.79] | 0.841 | 0.94[0.49-1.81] | 0.845 |
|  | Governmental employee | 84 | 13.5 | 44 | 7.1 | 2.44[1.51-3.93] | 0.000** | 1.74[0.85-3.58] | 0.128 |
|  | Private employee | 14 | 2.2 | 12 | 1.9 | 1.49[0.65-3.42] | 0.346 | 0.97[0.33-2.80] | 0.951 |
|  | Un employee | 29 | 4.7 | 9 | 1.7 | 0.40[0.18-0.89] | 0.025* | 0.53[0.19-1.51] | 0.234 |
| Source of information | Media | 71 | 12.5 | 114 | 19.9 | 1 |  | 1 |  |
|  | Health worker | 71 | 12.5 | 45 | 7.9 | 2.53[1.57-4.08] | <0.001** | 1.87[1.04-3.34] | 0.036** |
|  | Friends/relatives | 148 | 26.1 | 119 | 21 | 1.99[1.36-2.92] | <0.001** | 1.65[1.04-2.62] | 0.034* |
| Friend or relative with DM | I don't know | 8 | 1.3 | 44 | 7.1 | 1 |  | 1 |  |
|  | No | 137 | 22 | 258 | 41.4 | 1.86[0.83-4.19] | 0.135 | 1.58[0.66-3.79] | 0.303 |
|  | Yes | 148 | 23.2 | 28 | 4.5 | 11.77[5.01-27.68] | 0.000** | 4.03[1.56-10.47] | 0.004* |
| Family history of DM | No | 172 | 27.6 | 317 | 50.9 | 1 |  | 1 |  |
|  | Yes | 121 | 19.4 | 13 | 2.1 | 17.15[9.40-31.30] | 0.000** | 9.47[4.74-18.90] | 0.000** |
| ** Significant predictable variable with statistically significant at p<0.001. * Statistical significant with p Value <0.05. |  |  |  |  |  |  |  |  |  |

## 6. Discussion

This community-based cross-sectional study was conducted in Arba Minch town to determine the knowledge of BRF and associated factors regarding T2DM among WRA.

According to the results obtained from this study, 293 (47.0%) [95% of CI, 43.5-50.9] participants had good knowledge of BRF of T2DM. This finding was consistent with studies conducted in Ethiopia, Bale (48%), Nepal (48%), China (48.3%) and Tanzania (48.9%) respectively (19-22). The possible reason might be due to the fact that a number of participants heard about T2DM from their friends or relatives around the locations where the survey was conducted. Besides this, it was lower than the knowledge level reported in the studies done in Iran (23), Pakistan (24), Poland (25), Malaysia (26, 27), Nigeria (16) and Debrebrhan (28). This might be due to less information dissemination concerning T2DM and the difference in the study population in which the study was conducted among subjects with diabetes and their family members. On the other hand, this result is higher than the study conducted in Stalowa Wola, Poland [30%] (29), Saudi Arabia 29.6% -42.6 % (30, 31), 25.4 % in rural Tanzania (61), 19.6% in south western Nigeria (15), Indian western Rajasthan [17.6%] (32), Bangladesh [13%] (18), 17.3% in Mekele and 5 % in West Go jam (33, 34), Nepal [5.5-20%] (21, 35), and Japan [2.47%] (36). This inconsistency might be due to the sociocultural difference between the populations.

In comparison to study participants with average family incomes of less than 1999 Eth. birr, those with average family incomes between 3000 and 5000 Eth. birr and above 5001 Eth. birr had odds of having good knowledge that were 1.81 and 1.93 times higher, respectively. This may be because those with a higher income will have access to necessary information about the disease. The higher the income level, the higher the knowledge level. This study is incomparable with the studies done in Pakistan (24) and Bale Goba (19). This difference may be related to different study setups and socio-economic status. However, it was not found to be significantly associated in other south Indian studies (37).

In this study, more than half of the study participants (53.0 %) had poor knowledge of BRF of T2DM. The knowledge gap in the WRA may worsen the burden of the illness. This also indicates the need to encourage public health. The odds of good knowledge regarding BRF of T2DM among study participants who had a family history of DM were 9.47 times greater than the odds of knowledge with no DM in the family. Fortunately, this may be due to the fact that individuals with a family history of DM may develop a sense of vulnerability and susceptibility, which may strengthen their knowledge about the disease. This finding is also congruent with studies done in Mekele (33), New York (17), Bangladesh (18), Saudi Arabia (30), Nepal (35), Malaysia (27), and South India (37).

In this study, having a friend or relative history of DM had an association with knowledge which is similar with the Jordan (38) and china (20) studies. A friend or relative with DM was four times more likely to have knowledge than their counterparts. This could be due to a strong and intimate bond with DM friends or relatives who freely discuss their illness and those individuals with DM are not hesitant to disclose specific information about their disease with their friends.

The respondents’ diabetes information came from a variety of sources. Those who got information from a health worker were 1.87 times more likely to have good knowledge than those who got it from the media, and those who got it from friends or relatives were 65% more likely to have good knowledge. The information obtained from health workers may be more detailed, which is important for increasing individual understanding about BRF. Meanwhile, gathering information from friends and relatives in an intimate setting may aid them in persuasion and keep all relevant facts. This finding is consistent with a previous study in China (20). A study conducted in Bangladesh, however, it was not found to be significantly linked to good diabetes knowledge (18). This could be due to a lack of interest or that they did not seek any information about diseases.

This study revealed no association between BRF knowledge and occupation. A similar conclusion was made by a Nepalese author studying pregnant women (35). On the other hand, this result is incomparable to Bangladesh’s (18) and Vietnam’s (39). Employees may have had the ability to communicate scientific information with their colleagues and staff members, making them more knowledgeable than housewives, who were more focused to caring for their household.

Age had no significant relationship with behavioral risk knowledge in this survey. Similarly, in other studies, it had no association in Malaysia (27). The possible reason may be because of having different age groups. whereas, unlike in this study, age is a significant factor in the studies of Saudi Arabia (31) and Vietnam (39). This is probably due to less variation in age group in the study population.

Although there was no association between knowledge and educational status in this study, higher educational status was a major related factor in Saudi Arabia (31, 40), South India (37), and Jordan (38). Fortunately, this could be for the reason that people with a higher educational level pay more attention to information.

## 7. Conclusion

Overall, comparing with previous study regarding BRF of T2DM, the level of good knowledge was less than half of the participants. There was a statistically significant association between average family incomes, family history of diabetes, DM in the friend or relatives, and sources of information with the knowledge level. Nearly 60% of the respondents had no idea about others T2DM risk factors: pregnancy (gestational diabetes), sedentary life and sleeplessness that could liable to diabetes. A poor score in these areas could indicate a knowledge gap.

T2DM can be prevented with lifestyles modification through educating people to raise awareness concerning the risk factors. Hence, Diabetic educational platforms could play a pivotal role in controlling of diabetes. As a result, community health education programs about DM risk factors are required. Women, in particular reproductive age group, must have an in-depth understanding of BRF in order to improve their health and prevent the condition in the future.

Those who consume fattier and sweets on a daily basis and do not participate in maintaining a healthy weight may develop another risk, which can lead to T2DM. Surprisingly, in this study, those who checked their blood sugar and blood pressure frequently were less than 5%. This might indicate that women are ignoring their health. To better manage all women demand equal access to information. Consequently, knowledge could aid in assessing diabetes risk. Community-based education program about T2DM risk factors should also be conducted among the people at regular interval of time so as to make their knowledge at high level. To summarize, education on diabetes and socio demographic factors must be observed in order to improve their understanding on T2DM risk factors.

## Data Availability

All relevant data are within the manuscript and its Supporting Information files.

## Acknowledgement

Authors would like to thank Arba Minch collage of health science, for initiating to conduct this study. Deep appreciations have gone to Arba Minch general hospital, administration office, Arba Minch town health office and all administrators of the selected kebeles which were included in the study. Special thanks to all respondents, data collectors, supervisors and all peoples who involved in the study directly or indirectly.

## Supporting information

S1 fig. **The level of knowledge of Behavioral Risk Factors for Type-2DM among reproductive age women in Arba Minch town, 2022(n=623)**.

S2 fig. **The response of reproductive age women on their knowledge of symptoms for T2DM in Arba Minch, 2022(N=623)**.

S1Table. **Socio-demographic status of respondents on Behavioral Risk Factors of Type 2-Diabetes Mellitus in Arba Minch town 2022(n=623)**.

S2 table. **The description of response of study participant on the socio-economic and awareness status (n=623)**.

S3 table. **The description of general knowledge about Type-2-DM definition among Reproductive age Women in Arba Minch, 2022(n=623)**.

S4 table. **The Frequency distribution of participants response on knowledge of Behavioral risk factors for type 2 diabetes, Arba Minch town, 2022 (n= 623)**

S5 table. **The knowledge of prevention of Type 2DM and its complication among Reproductive Age Women in Arba Minch town, 2022(n=623)**.

S6 table. **The perception of reproductive age women on behavioral risks of Type 2DM in Arba Minch town, 2022(n=623)**.

S7 table. **The description of behavioural risk and adoption of health status of Women Reproductive Age for Type 2DM in Arba Minch town 2022(n=623)**

S8 table. **Bi-variable and multi-variable logistic regression predicting knowledge related to Behavioral Risk Factors for Type 2 Diabetes Mellitus among women reproductive age group of Gamo zone, Arba Minch towns, 2022 (N = 623)**.

